# Comparative Evaluation of Blood Pressure, Central Adiposity, and Physical Activity Patterns Between Wheelchair Users and Ambulatory Adults in a Rural South African Settings: A Cross-Sectional Study Protocol

**DOI:** 10.64898/2026.09.16.26363280

**Authors:** Thendo Nefale, Musa L. Mathunjwa, Brandon S Shaw, Gudani G. Mukoma

## Abstract

**Background:** Hypertension is a major modifiable driver of cardiovascular disease in low- and middle-income countries, heavily driven by central adiposity and physical inactivity. Wheelchair users represent a structurally marginalized population facing heightened exposure to prolonged sitting and altered body composition. However, context-specific epidemiological data comparing cardiovascular risk profiles by mobility status in rural South Africa remain scarce.

**Objective:** This study aims to compare blood pressure profiles, central adiposity (waist circumference), physical activity levels, and sedentary behavior between wheelchair users and ambulatory adults in the Thulamela Municipality, Limpopo Province, and to determine if mobility status is independently associated with hypertension.

**Methods:** This study utilizes an analytical, community-based cross-sectional design. A sample of N = 210 adults (n = 70 wheelchair users; n = 140 ambulatory controls) will be recruited using purposive and convenience sampling strategies within rural networks. Assessments will feature seated blood pressure measurements and seated waist circumference metrics to standardize data collection across both groups. Physical activity and sedentary durations will be quantified using the Leisure Time Physical Activity Questionnaire for Spinal Cord Injury (LTPAQ-SCI) and the International Physical Activity Questionnaire Short Form (IPAQ-SF). Outcomes will be mathematically harmonized into weekly minutes of moderate-to-vigorous physical activity (MVPA).

**Analysis:** Inter-group variations will be assessed using independent samples t-tests or Mann-Whitney U tests. Multivariate binary logistic regression models will isolate the independent association between mobility status and hypertension, controlling for confounding clinical and demographic factors.

**Strengths and Limitations of This Study:**

- **Strength:** Standardizes both hemodynamic (blood pressure) and anthropometric (waist circumference) measurements in a seated position to eliminate positional bias between wheelchair and ambulatory cohorts.
- **Strength:** Harmonizes two population-validated physical activity questionnaires (LTPAQ-SCI and IPAQ-SF) into a single continuous metric (MVPA) to bridge functional measurement gaps.
- **Strength:** Targets an epidemiologically under-researched, structurally marginalized population in rural, resource-constrained South African settings.
- **Limitation:** The cross-sectional design limits the ability to establish causal relationships between mobility status, central adiposity, and the onset of hypertension.
- **Limitation:** Relying on self-reported questionnaires for physical activity and sedentary behavior introduces potential recall and social desirability biases compared to device-based accelerometry.

## Introduction

Globally, cardiovascular disease (CVD) remains the leading cause of mortality, with hypertension acting as its most pervasive modifiable driver, affecting approximately 1.4 billion adults worldwide (WHO, 2025). Over two-thirds of this burden is concentrated in low- and middle-income countries, with Sub-Saharan Africa facing a rising pooled prevalence of 27% driven by rapid epidemiological shifts, including urbanization and sedentary lifestyles (Kakoma et al., 2026). Nationally, South Africa is facing a severe non-communicable disease crisis. Recent data indicates that hypertension affects roughly 45% of men and 48% of women over the age of 15, with over 8.2 million citizens currently diagnosed (National Department of Health (NDoH), 2019; Sobuwa, 2025). This escalating trend heavily impacts rural, resource-constrained environments where systemic health inequities, high unemployment, and poor access to primary healthcare worsen secondary metabolic risk factors, such as central adiposity and severe physical inactivity.

Among vulnerable demographics, wheelchair users represent a highly marginalized population with uniquely elevated cardiometabolic vulnerabilities (Andrabi et al., 2022a). The physiological intersection of prolonged, structurally forced sitting and limited lower-limb locomotion profoundly impacts body composition, precipitating skeletal muscle atrophy and increased visceral adipose tissue accumulation (de Groot et al., 2024). Consequently, individuals with mobility impairments are over 1.5 times more likely to experience clinical CVD events than their ambulatory peers (Andrabi et al., 2022b; Mathunjwa et al., 2023).

Despite these clear dangers, a major epidemiological gap persists. Most domestic public health surveillance models and cardiovascular risk profiles are derived exclusively from ambulatory cohorts. As a result, there is a scarce amount of comparative data quantifying how mobility limitations independently compound blood pressure elevation and central adiposity within rural African contexts, such as the Thulamela Municipality. This lack of evidence prevents public health systems from designing targeted, accessible, and inclusive preventative healthcare initiatives.

To bridge this gap, this study aims to compare blood pressure profiles, central adiposity (waist circumference), physical activity levels, and sedentary behavior between wheelchair users and ambulatory adults in the Thulamela Municipality, Limpopo Province. Furthermore, it seeks to determine if mobility status is independently associated with hypertension after controlling for key clinical and socio-demographic covariates.

## 2. Methodology

### 2.1 Research Design and Setting

This study will employ an analytical, community-based cross-sectional research design. The study will be conducted within the rural communities and primary health care clinics of the Thulamela Local Municipality, located in the Vhembe District of the Limpopo Province, South Africa. This geographic region is characterized by resource-constrained settings, structural infrastructure challenges, and limited access to specialized physical rehabilitation facilities (Thulamela Local Municipality, 2016).

### 2.2 Participant Sampling and Recruitment

A total sample of *N* = 210 participants will be recruited using a combination of purposive and convenience sampling techniques through local community disability forums, clinic registries, and traditional leadership networks. The sample will be stratified into two comparative cohorts based on an allocation ratio of 1:2.

#### Wheelchair Users (*n* = 70)

Adults aged ≥ 18years who have a permanent lower-limb mobility impairment and have relied primarily on wheelchair for daily locomotion for at least 6 months. Individuals with active pressure sores, acute systemic infections, or terminal illnesses will be excluded.

#### Ambulatory Controls (*n* = 140)

Age- and sex-matched adults living in the same socio-geographic neighborhoods who are able to walk independently without assistive devices.

### 2.3 Data Collection Procedures and Instrumentation

Data collection will be conducted by trained research assistants at localized community centers or primary care facilities. To ensure consistency, all physical measurements for both groups will be adapted to a standardized seated position.

#### 2.3.1 Hemodynamic Assessment (Blood Pressure)

Blood pressure (BP) will be measured using a validated, automated digital sphygmomanometer fitted with an appropriate cuff size. Participants will rest quietly in a seated position for 5 minutes prior to testing. Two separate readings will be taken from the left arm, separated by a 2-minute interval. If the variance between the two readings exceeds 10 mmHg for either systolic or diastolic BP, a third measurement will be taken. The average of the final two recorded measurements will be used for clinical classification (hypertension defined as systolic 130 mmHg and/or diastolic 80 mmHg, or current use of antihypertensive medication) following the American College of Sports Medicine (ACSM) guidelines.

#### 2.3.2 Central Adiposity (Waist Circumference)

To prevent positional bias between the cohorts, central adiposity will be operationalized via seated waist circumference (WC). Using a non-stretchable, flexible anthropometric tape measure, the measurement will be taken at the horizontal plane corresponding to the midpoint between the lowest rib margin and the superior border of the iliac crest. The measurement will be recorded to the nearest 0.1 cm at the end of a normal, unforced expiration.

#### 2.3.3 Physical Activity and Sedentary Behaviour

Due to the distinct mechanical profiles of locomotion, self-reported physical activity will be captured using population-specific instruments:

- **Wheelchair Cohort:** The Leisure Time Physical Activity Questionnaire for Spinal Cord Injury (LTPAQ-SCI) will be administered to quantify the frequency and duration (minutes/week) of mild, moderate, and heavy intensity physical activity.
- **Ambulatory Cohort:** The International Physical Activity Questionnaire Short Form (IPAQ-SF) will be utilized to capture weekly durations spent walking, as well as engaging in moderate-to-vigorous physical activity.

### 2.4 Proposed Study Timeline

The proposed study timeline is presented in Table 1 and outlines the anticipated sequence of proposal development, ethical approval, participant recruitment, data collection, data analysis, dissertation writing, and final submission. All activities will be subject to the required institutional ethics approval and permission from the relevant provincial health authorities.

**Table 1.** Proposed Study Timeline.

| Activities | Apr–Jun 2026 | Jun–Jul 2026 | Jul–Sep 2026 | Oct–Nov 2026 | Jan–Feb 2027 | Mar–May 2027 | Jun–Jul 2027 |
| --- | --- | --- | --- | --- | --- | --- | --- |
| Proposal development | ✓ |  |  |  |  |  |  |
| Literature review | ✓ | ✓ |  |  |  |  |  |
| Proposal submission and approval |  | ✓ | ✓ |  |  |  |  |
| Ethics submission and approval |  |  | ✓ | ✓ | ✓ |  |  |
| Pilot testing of tools |  |  |  |  | ✓ |  |  |
| Participant recruitment |  |  |  |  |  | ✓ |  |
| Data collection |  |  |  |  |  | ✓ |  |
| Data capturing and cleaning |  |  |  |  |  | ✓ | ✓ |
| Data analysis |  |  |  |  |  |  | ✓ |
| Results interpretation |  |  |  |  |  |  | ✓ |
| Dissertation writing |  |  |  |  |  |  | ✓ |
| Editing and final submission |  |  |  |  |  |  | ✓ |

Note: Participant recruitment and data collection are planned for 1 March–31 May 2027, subject to ethics approval and permission from the relevant health authorities.

### 2.5 Statistical Power and Sample Size

The sample size estimation was executed using *G* * Powersoftware (version 3.1). Setting a moderate Cohen’s effect size (*d* = 0.50), an alpha level of *α* = 0.05, a target statistical power of 1 ― *β* = 0.80, and an allocation ratio of 1:2, the calculated minimum sample size was determined to be *N* = 192(*n* = 64 wheelchair users; *n* = 128ambulatory controls). Adjusting for a potential 10%missing data threshold, the final target recruitment cohort is established at *N* = 210.

## 3.

**Table 2.** Data Collection Protocols.

| Measurement Category | Primary Tool/Instrument | Procedural Specifics |
| --- | --- | --- |
| Hemodynamic Assessment | Validated Automated Sphygmomanometer | Seated posture, 2 separate readings. If variance exceeds >10 mmHg a 3rd reading is taken and the final two are averaged. Classified via ACSM guidelines. |
| Central Adiposity | Non-stretchable, inflexible tape measure | Standardized seated posture for both groups to avoid positional bias. Midpoint between lower rib margin and iliac crest, measured at the end of a normal expiration. |
| Wheelchair Physical Activity | LTPAQ-SCI questionnaire | Quantifies weekly minutes of mild, moderate, and heavy leisure-time physical activity specific to wheelchair mechanics |
| Ambulatory Physical Activity | IPAQ-SF questionnaire | Documents physical activity duration across moderate and vigorous domains for walking controls. |
| Sedentary Behavior | Standardized Self-Report Survey | Captures total daily sitting time in hours/day to evaluate resting exposures. |

Methodological Note on Physical Activity Harmonization: To allow statistical comparison, physical activity data from the LTPAQ-SCI and IPAQ-SF will be mathematically harmonized into a single continuous vector representing weekly minutes of moderate-to-vigorous physical activity (MVPA). Data interpretations will take into account the distinct domain variations inherent to each original instrument.

## 4. Statistical Analysis Plan

Statistical processing will be performed using IBM SPSS Statistics software, with the minimum threshold for significance set at *p* < 0.05. Normality distributions will be mapped via Shapiro-Wilk testing.

- **Comparative Unimodal Analyses:** Independent samples *t*-tests (parametric) or Mann-Whitney *U*tests (non-parametric) will be utilized to contrast continuous blood pressure readings and waist circumferences between cohorts. Proportional hypertension rates will be compared using Chi-square tests (*Χ*^2^).
- **Multivariate Modeling:** A robust binary logistic regression architecture will be constructed to evaluate the primary research question. The dependent variable is dichotomized as hypertension status (hypertensive vs. non-hypertensive). Mobility status (wheelchair vs. ambulatory) will serve as the primary independent predictor, adjusting for known clinical and demographic covariates, including age, sex, waist circumference, cumulative MVPA minutes, sedentary duration, smoking habits, and alcohol consumption patterns. Adjusted Odds Ratios (AOR) accompanied by 95% confidence intervals will be reported.

### Missing Data Management Strategy

Although sample size computations account for a 10% missing data buffer, any actual incomplete data points across variables will be analyzed for patterns. If data are found to be Missing Completely at Random (MCAR) or Missing at Random (MAR), a complete-case analysis approach will be utilized for minor losses (<5%). For primary outcomes exceeding this threshold, multiple imputation procedures using chained equations will be deployed to protect statistical power and minimize parameter bias.

## 5. Discussion

The primary objective of this study is to compare blood pressure profiles, central adiposity, and physical activity patterns between wheelchair users and ambulatory controls within a rural South African community. This protocol addresses a significant gap in regional epidemiological data, as public health surveillance in Sub-Saharan Africa often overlooks populations with physical disabilities.

Physiologically, the prolonged sitting required by wheelchair locomotion leads to localized muscular inactivity and a reduction in daily energy expenditure (WHO, 2020). These factors contribute to changes in body composition, specifically muscle atrophy and increased visceral fat accumulation, which can be measured via seated waist circumference (Kim et al., 2024; Sugiyama and Furukawa, 2026). These changes often drive metabolic issues and increase arterial stiffness, predisposing wheelchair users to chronic hypertension (Kim et al., 2024). By using a comparative cross-sectional design, this study can clarify how much mobility limitations independently increase cardiovascular risk beyond traditional lifestyle factors (Dube et al., 2025; Mathunjwa et al., 2023).

A notable feature of this protocol is adapting the data collection methods to fit the physical constraints of wheelchair users. Measuring waist circumference and blood pressure in a standardized seated position minimizes positional errors and allows for a direct comparison with ambulatory controls (Sumrell et al., 2018; Muntner et al., 2019). Additionally, mathematically combining data from the LTPAQ-SCI and the IPAQ-SF into weekly minutes of moderate-to-vigorous physical activity (MVPA) helps reconcile the functional differences between wheelchair use and upright walking (Martin Ginis et al., 2012; WHO, 2020).

Conducting this research in the Thulamela Municipality highlights how environmental and socio-economic factors shape physical activity. In rural South Africa, challenges like uneven terrain, lack of accessible transport, and limited community infrastructure create major barriers to active living for individuals with physical impairments (van Biljon et al., 2022; Douglas et al., 2021). The results of this study will provide essential evidence to help local public health networks design more inclusive preventive healthcare programs and adaptive physical activity interventions.

## 6. Patient and Public Involvement (PPI)

Neither patients nor the broader rural disability public were directly involved in formulating the underlying core epidemiological research questions, deciding the cross-sectional stratification allocation ratios, or drafting the specific choices of anthropometric metrics. However, during the preparatory phases, leaders from local community disability forums and traditional networks will be consulted to assess the operational feasibility of localized testing centers, ensure the cultural appropriateness of the bilingual (English/Tshivenda) tool translations, and review the physical participant burden associated with the seated testing procedures.

## 7. Data Management and Quality Control

To guarantee data integrity, all primary source data collected via case report forms and self-report surveys will be processed using centralized electronic data capture software configured with real-time range checks, data-type validations, and missingness prompts to mitigate human data entry errors. All data assets will be completely de-identified through assigned alphanumeric participant numbers. The electronic master database will be stored on password-protected cloud servers managed by institutional infrastructure, incorporating multi-factor authentication. Access will be strictly confined to authorized investigators. In compliance with local ethical governance frameworks, raw and processed scientific data vectors will be safely retained for a mandatory minimum curation period of 10 years before destruction.

## Data Availability

No datasets were generated or analysed during the current study. All relevant data generated from this study will be made available upon study completion, subject to applicable ethical and data-protection requirements.

## 8. Ethical Considerations & Governance

### 8.1 Institutional Oversight

Ethical oversight is provided by the institutional research ethics committee. Fully informed, written consent will be secured from all candidates prior to testing, utilizing bilingual information sheets and declarations available in both English and Tshivenda. Because testing protocols (blood pressure and tape measurements) are non-invasive, physical risk exposures are minimized. Standardized participant numbers will be used to protect personal data throughout processing and secure storage.

### 8.2 Ethics Approval and Consent to Participate

This study protocol will be reviewed and approved by the Institutional Research Ethics Committee (IREC) of the participating institution and relevant provincial health authorities before implementation. The study will be conducted in strict accordance with the Declaration of Helsinki. Fully informed, voluntary, written consent will be obtained from all participants prior to the commencement of any data collection procedures. Information sheets and informed consent forms will be provided in both English and Tshivenda to accommodate local linguistic preferences. Participants will be explicitly informed of their right to withdraw from the study at any stage without facing any negative consequences. Ethics status: The study will not commence until written approval has been granted by the relevant institutional research ethics committee and permission has been obtained from the relevant provincial health authorities.

### 8.3 Author Contributions

All authors contributed significantly to the conceptualization and structural design of this study protocol: TN drafted the primary study protocol manuscript, developed the operational data collection methodologies, and designed the physical activity harmonization framework. MLM provided oversight on the study design, refined the clinical hemodynamic and anthropometric assessment procedures, and structured the multivariate statistical analysis plan. GGM contributed to the contextual framework of the study, managed local logistical coordination for the rural research setting, and critically revised the manuscript for scientific clarity. All authors have read, reviewed, and formally approved the final version of this manuscript before submission.

### 8.5 Data Availability Statement

No datasets have been generated or analysed at the time of protocol submission. Data sharing will be addressed in accordance with the approved ethics protocol and applicable institutional and legal requirements.

### 8.6. Funding

This research project received institutional support from the Department of Human Movement Science at the the participating institution. No external commercial funding or corporate sponsorships were associated with this study.

